# Longitudinal associations between stroke and psychosis: a population-based study

**DOI:** 10.1101/2022.11.22.22282626

**Authors:** Alvin Richards-Belle, Norman Poole, David P.J. Osborn, Vaughan Bell

## Abstract

**Background:** The co-occurrence of stroke and psychosis is a serious neuropsychiatric condition, but little is known about the course of this comorbidity.

**Aims:** To estimate longitudinal associations between stroke and psychosis over 10 years.

**Methods:** A 10-year population-based study using data from the English Longitudinal Study of Ageing. A structured health assessment recorded i) first-occurrence stroke and ii) psychosis, at each wave. Each were considered exposures and outcomes in separate analyses. Logistic and Cox proportional hazards regression and Kaplan-Meier methods were used. Models were adjusted for demographic and health behaviour covariates, with missing covariates imputed using random forest multiple imputation.

**Results:** Of 19,808 participants, 24 reported both stroke and psychosis (median Wave 1 age 63, 71% female, 50% lowest quintile of net financial wealth) at any point during follow-up. By 10 years, the probability of an incident first stroke in participants with psychosis was 21.4% (95% CI, 12.1 to 29.6) compared to 8.3% (95% CI, 7.8 to 8.8) in those without psychosis (absolute difference: 13.1%; 95% CI, 20.8 to 4.3, log rank *p*<0.001; fully-adjusted hazard ratio (HR): 3.63; 95% CI, 2.25 to 5.87). The probability of reporting incident psychosis in participants with stroke was 2.3% (95% CI, 1.4 to 3.2) compared to 0.9% (95% CI, 0.7 to 1.1) in those without (absolute difference: 1.4%; 95% CI, 0.7 to 2.1, log rank *p*<0.001; fully-adjusted HR: 5.81; 95% CI, 2.89 to 11.70).

**Conclusions:** Stroke is an independent predictor of psychosis (and vice versa), after adjustment for potential confounders.

## Introduction

Stroke and psychosis are both severe, disabling conditions, independently associated with a high risk of morbidity and mortality, meaning that their co-occurrence is considered to be a particularly serious neuropsychiatric condition.^1,2^ Clinically, people affected by both conditions are less likely to receive either adequate stroke or psychiatric care^3,4^ indicating a clear need to develop the evidence-base and clinical pathways. However, research on stroke-psychosis comorbidity is limited to cohort studies focusing on stroke risk in people with schizophrenia^5^ and bipolar disorder^6^ diagnoses, while the majority of publications on psychosis after stroke are single case studies.^7^

Although psychosis has been frequently described as a “rare” complication of stroke,^8^ a meta-analysis of post-stroke psychosis reported the prevalence of delusion and hallucination to be 4.67% and 5.05%, respectively.^9^ In terms of stroke occurring after psychosis (‘post-psychosis stroke’), a meta-analysis by Li et al reported that individuals with schizophrenia have an increased risk of stroke (relative risk 1.50; 95% confidence interval [CI] 1.25 to 1.80) and an increased risk of stroke mortality (relative risk 1.65; 95% CI, 1.31 to 2.08).^5^ These findings are echoed in individuals with bipolar disorder, where a recent meta-analysis by Yuan et al reported increased stroke incidence (hazard ratio 1.43; 95% CI, 1.09 to 2.18) and stroke mortality (hazard ratio 1.35; 95% CI, 1.26 to 1.45). A recent cross-sectional study that combined data from national representative psychiatric epidemiology studies from four countries (UK, USA, Chile and Colombia) reported that 3.81% of people (approximately 1 in 26) with probable psychosis have stroke, while 3.15% of people with stroke (approximately 1 in 32) have probable psychosis.^10^ These studies indicate a higher level of co-occurrence than has been assumed in, previous, admittedly ad-hoc estimates, of “rare” occurrence in the literature.

To date, however, estimates of risk and co-occurrence have been taken from studies that have either solely focused on stroke incidence in a psychosis population, or psychosis incidence in a stroke population. Methodologically, they have either been cross-sectional or cohort studies with limited waves of follow-up. This has meant it has been impossible to estimate how stroke-first or psychosis-first presentations differ in terms of risk for stroke-psychosis comorbidity - something important to estimate within the same population - or the extent to which the risk for comorbidity changes over time. Initial evidence suggests that risk of psychosis after stroke may peak at approximately six months post-stroke.^9^ Conversely, cardiovascular risk factors compound over time in people with psychosis,^11^ potentially leading to a late rather than ‘early peak’ risk profile. Indeed, Fleetwood et al reported this pattern in individuals with schizophrenia and bipolar disorder where stroke risk markedly increased at five year follow-up but remained similar between 30-day and 1-year follow-up.^12^

One challenge, however, is that few studies measure both psychosis and stroke within the same cohort, something needed for a direct comparison between time to onset. One exception is the English Longitudinal Study of Ageing (ELSA),^13^ a representative longitudinal study of adults aged 50 years and older that collects economic, social, psychological, cognitive, health, biological and genetic data which was initiated in 2002 and is ongoing. We used data from ELSA to estimate associations between stroke and psychosis and the incidence of stroke after psychosis and psychosis after stroke over 10 years of follow-up.

## Methods

### Study design

This population-based longitudinal study used data from the English Longitudinal Study of Ageing (ELSA). ELSA is an ongoing, prospective, observational, longitudinal study that began in 2002 and which includes a representative sample of adults aged 50 years and over in England and their cohabiting partners of any age (partners were not selected based on representativeness).^13^ ELSA received ethical approval from the London Multi-Centre Research Ethics Committee (REC) (waves 1-3, references: MREC/01/2/91, MREC/04/2006, 05/MRE02/63), National Hospital for Neurology and Neurosurgery & Institute of Neurology Joint REC (wave 4, reference: 07/H0716/48), Berkshire REC (wave 5, reference: 09/H0505/124), NRES Committee South Central – Berkshire (waves 6-9, references: 11/SC/0374, 13/SC/0532, 15/SC/0526, 17/SC/0588). All participants provided informed consent. ELSA data are publicly accessible; data were accessed via UK Data Service (project ID: 222747).

### Participants

Full details of ELSA methods are described in Steptoe et al.^13^ ELSA comprises ‘core members’ and their partners as participants. Core members, intended to be nationally representative, were sourced from their prior participation in the Health Survey for England (HSE) (a national survey monitoring health and care trends).^14^ The original core members, selected from the HSE carried out in 1998, 1999 and 2001, were eligible for ELSA if they agreed to follow-up during their HSE participation, were born prior to 1952, and lived in private accommodation. Currently, core members and their partners are followed-up every two years. Refresher samples of adults aged ≥50 years (and their partners) are added to the study periodically to ensure continuing representativeness. To ensure maximal sample size, we included ELSA participants (core members and partners) who took part in at least one interview data collection wave between 2002 and 2019 (ELSA waves 1 to 9). Trained interviewers carried out fieldwork interviews in participants’ homes (or institutions, if that was where a participant was now residing). Where necessary, interviews were carried out with proxies (e.g., if the participant lacked mental capacity or was a hospital inpatient during data collection).

### Outcomes and exposures

Stroke and psychosis were each considered as main outcomes and as exposures in separate analyses. At each wave, participants were asked if a doctor had ever diagnosed them with “a stroke (cerebral vascular disease)” as part of a structured health assessment. Free-text responses indicative of stroke were coded as stroke by the interviewer, where necessary.

Analysis of the relationship between self-report and medically verified strokes in ELSA’s sister study, the Health and Retirement Study, replicated known risk factor associations and indicated misreporting was non-systematic, indicating that participant-report can be used to study stroke risk.^15^ However, due to the longitudinal nature of the study, we added a second validation step by checking that earlier wave self-reported strokes were also self-reported in later waves. If there was a discrepancy between an earlier self-reported stroke and a later self-report, then their stroke data for the previous wave was recoded to no stroke. Data on age at first stroke and number of stroke recurrences (0, 1, 2, or 3+) were also reported; age at first stroke was calculated as the youngest reported stroke age, and number of stroke recurrences calculated as the sum of total number of reported stroke recurrences, across all waves.

Strokes (“stroke/cerebral haemorrhage/cerebral thrombosis”) reported by participants in the HSE, prior to their ELSA participation, were also included in this study (this data was available only for participants who joined ELSA from HSE survey years between 1998 to 2006).

At each ELSA wave, participants were also asked if a doctor had ever diagnosed them with “any emotional, nervous or psychiatric problems.” Participants responding affirmatively to this screening question were then probed to identify which of the following problems they had; hallucinations, anxiety, depression, emotional problems, schizophrenia, psychosis, mood swings, and/or manic depression. Psychosis was defined in this study as either a report of “hallucinations”, “schizophrenia” or “psychosis”.

### Covariates

In addition to unadjusted analyses, two adjusted models were reported. The first (model 1) adjusted for demographic characteristics; age at wave 1 (calculated by subtracting the year of birth from 2002), sex, ethnicity (white compared to other ethnicities) and baseline net financial wealth. Where a participant had multiple ethnicities recorded across waves, the most frequent was used (or the most recent, if frequencies were equal). Quintile of net financial wealth, defined as participants’ gross financial wealth minus debt, was included as a proxy for socioeconomic status (as is common in studies of older adults and consistent with a previous study using ELSA data).^16^ The second models (model 2) additionally adjusted for baseline cigarette smoking status, level of vigorous physical activity, and level of alcohol consumption. Smoking status referred to whether the participant *currently* smoked at the time of data collection. Frequency of vigorous physical activity, in reference to sports and activities in daily life, was considered according to the following levels: ‘hardly ever or never’, ‘1-3 times a month’, ‘once a week’ or ‘more than once a week’. Frequency of alcohol consumption referred to the last 12 months; different responses scales were used across waves, these were recoded to ‘daily/almost daily’, ‘1-4 times/week’, ‘monthly’, ‘rarely/special occasions only’ or ‘not at all’ for consistency.

In addition to outcomes/exposures and covariates, participants were described in terms of ever-reporting depression or anxiety across waves 1-9, study participation (i.e., number of waves participated, whether took part in all waves) and whether participants died prior to wave 6 (mortality status was well characterised only up to this time-point).

### Analysis

Data were anchored to the first wave that participants took part in (e.g., if a participant joined ELSA at wave 4, then wave 4 was taken as their baseline). Means with standard deviations (SD), medians with interquartile ranges (IQR), and/or counts and proportions were used for descriptive statistics, as appropriate. Follow-up times were converted from number of waves to years for interpretability.

Logistic regression was used to investigate the associations between stroke and psychosis, with unadjusted and adjusted (see *Covariates*) odds ratios (OR) and associated 95% CIs, reported after 4 and 10 years of follow-up. Kaplan-Meier probability estimates (with 95% CIs) and survival curves were used to summarise and illustrate the cumulative incidence of first-reported stroke in participants with and without psychosis up to 10 years. The log-rank test was used to test differences in being stroke-free at the end of follow-up between the groups. Cox proportional hazards regression was used to estimate the association between psychosis on the hazard of stroke; unadjusted and adjusted (see *Covariates*) hazard ratios (HR) with 95% CIs were reported. In participants reporting psychosis, the start of follow-up was defined as the wave that they reported psychosis for the first time. For those not reporting psychosis, the start of follow-up was the first wave that they participated in. The end of follow-up was defined as either the wave that a stroke was first reported (in those reporting a stroke) or the last wave that the participant took part in (in those not reporting stroke). Follow-up time was set to zero if first stroke was reported prior to psychosis and if stroke and psychosis were first reported during the same wave. The same methods were used to investigate the effect of stroke on incidence/hazard of stroke. All adjusted survival models included a shared frailty term to account for individual random effects.

Adjusted results are reported following multiple imputation as well as from complete-case analysis. Random forest multiple imputation, implemented using the *missForest* package in R, was used to impute values for missing baseline covariates (outcomes were not imputed). This method was used as it allows for simultaneous imputation of numeric and categorical variables, it does not rely on distributional assumptions - allowing for complex interactions and non-linear relations between variables, and has been shown to outperform several other imputation methods.^17^

Analyses were conducted using *R* version 4.0.3^18^ and the full code and output for the analysis is available in the format of a Jupyter Notebook,^19^ a document that combines both code and the output in a form that can be re-run and reproduced. All code and output is available on the online archive: https://github.com/vaughanbell/longitudinal-stroke-psychosis-ELSA

## Results

### Participants

Demographics and descriptive statistics for the sample are shown in Table 1. Of 19,808 participants who took part in at least one wave between 2002 and 2019, a total of 1,279 (6.5%) reported stroke and 150 (0.8%) reported psychosis at least once. Twenty-four participants reported both stroke and psychosis at any point across waves - equating to 1.9% of the stroke population and 16.0% of the psychosis population.

**Table 1.** Demographics and descriptive statistics for sample.

|  | *No Stroke or*<br>*Psychosis* | *Psychosis only* | *Stroke and*<br>*Psychosis* | *Stroke only* |
| --- | --- | --- | --- | --- |
| N | 18,403 | 126 | 24 | 1,255 |
| Age at Wave 1<sup>1</sup> | 56 (48, 66) | 52 (47, 58) | 63 (57, 77) | 69 (60, 77) |
| Age categories, n (%) |  |  |  |  |
| <60 | 11,063 (60.1%) | 97 (77.0%) | 9 (37.5%) | 296 (23.6%) |
| 60-69 | 3,913 (21.3%) | 16 (12.7%) | 5 (20.8%) | 346 (27.6%) |
| 70+ | 3,425 (18.6%) | 13 (10.3%) | 10 (41.7%) | 613 (48.8%) |
| Unknown | 2 | 0 | 0 | 0 |
| Sex |  |  |  |  |
| Male | 8,307 (45.1%) | 56 (44.4%) | 7 (29.2%) | 641 (51.1%) |
| Female | 10,096 (54.9%) | 70 (55.6%) | 17 (70.8%) | 614 (48.9%) |
| Ethnicity |  |  |  |  |
| White | 16,706 (90.8%) | 116 (92.1%) | 22 (91.7%) | 1,128 (89.9%) |
| Other | 799 (4.3%) | 5 (4.0%) | 0 (0.0%) | 40 (3.2%) |
| Unknown | 898 (4.9%) | 5 (4.0%) | 2 (8.3%) | 87 (6.9%) |
| Net financial wealth (quintile), n (%) |  |  |  |  |
| 1 (lowest) | 3,398 (22.4%) | 52 (46.8%) | 11 (50.0%) | 296 (25.5%) |
| 2 | 2,837 (18.7%) | 18 (16.2%) | 1 (4.5%) | 289 (24.8%) |
| 3 | 2,857 (18.9%) | 17 (15.3%) | 1 (4.5%) | 238 (20.5%) |
| 4 | 2,994 (19.8%) | 15 (13.5%) | 4 (18.2%) | 182 (15.6%) |
| 5 (highest) | 3,052 (20.2%) | 9 (8.1%) | 5 (22.7%) | 158 (13.6%) |
| Unknown | 3,265 | 15 | 2 | 92 |
| Smoked cigarettes at baseline, n (%) | 3,257 (18.3%) | 46 (36.8%) | 4 (16.7%) | 236 (18.8%) |
| Unknown | 601 | 1 | 0 | 3 |
| Level of vigorous activity at baseline, n (%) |  |  |  |  |
| Hardly ever or never | 10,499 (57.6%) | 99 (79.2%) | 17 (85.0%) | 932 (76.7%) |
| 1-3 times/month | 1,760 (9.7%) | 9 (7.2%) | 1 (5.0%) | 70 (5.8%) |
| Once a week | 1,861 (10.2%) | 7 (5.6%) | 1 (5.0%) | 75 (6.2%) |
| More than once a week | 4,093 (22.5%) | 10 (8.0%) | 1 (5.0%) | 138 (11.4%) |
| Unknown | 190 | 1 | 4 | 40 |
| Alcohol consumption at baseline, n (%) |  |  |  |  |
| Daily/almost daily | 4,189 (25.4%) | 18 (16.1%) | 5 (27.8%) | 281 (24.6%) |
| 1-4 times/week | 5,833 (35.4%) | 31 (27.7%) | 4 (22.2%) | 302 (26.4%) |
| Monthly | 1,801 (10.9%) | 11 (9.8%) | 4 (22.2%) | 134 (11.7%) |
| Rarely/special occasions only | 2,911 (17.7%) | 28 (25.0%) | 4 (22.2%) | 227 (19.9%) |
| Not at all | 1,732 (10.5%) | 24 (21.4%) | 1 (5.6%) | 198 (17.3%) |
| Unknown | 1,937 | 14 | 6 | 113 |

<sup>1</sup> Median (IQR)

Participants reporting both stroke and psychosis tended to be older than those reporting only psychosis but younger than those reporting only stroke. Almost three quarters (71%) of the stroke and psychosis group were female and half (50%) were in the lowest quintile of net financial wealth (double the proportion of those reporting just stroke or no stroke/psychosis at all). This group also had the lowest levels of baseline vigorous physical activity and highest level of alcohol use in the last 12 months, whilst the psychosis only group had the highest proportion of cigarette smokers. A high proportion of participants reporting psychosis also reported depression and anxiety, whereas the stroke only group were more similar to those without stroke or psychosis in this regard (Supplementary Table 1). Compared to those reporting stroke only, first stroke tended to occur earlier in participants who reported both stroke and psychosis (median age, 66 vs 69) and the mean total number of stroke recurrences was higher (2.2 vs 1.1) (Supplementary Table 1).

The rate of follow-up across waves was high – participants with stroke and psychosis or psychosis only participated in a median of six (out of nine) waves, compared with five waves for those reporting stroke only (lower likely due to the higher mortality rate in the latter group) (Supplementary Table 2).

### Stroke risk after psychosis

After 4 years of follow-up, the odds of stroke in people reporting psychosis were higher than in those without psychosis (unadjusted OR, 3.99, 95% CI, 2.39 to 6.34) (Table 2). Following multiple imputation to impute missing covariates (missing data is illustrated in Table 1) and adjustment, the ORs increased, and was highest in model 1 (adjusted for age, sex, ethnicity, and net financial wealth) (OR 5.52, 95% CI, 3.23 to 9.03). By 10 years, the ORs had reduced slightly, but psychosis was still significantly associated with an increased risk of stroke – resulting in an OR of 3.21 (95% CI, 1.97 to 5.00) in the model additionally adjusting for smoking status, level of vigorous physical activity and level of alcohol use (model 2).

**Table 2.** Odds ratios estimates for stroke risk after psychosis and psychosis risk after stroke.

|  | *Unadjusted*<br>*OR (95% CI)* | *Adjusted, model 1*<br>*OR (95% CI)* | *Adjusted, model 2*<br>*OR (95% CI)* |
| --- | --- | --- | --- |
| *Odds of stroke in psychosis* |  |  |  |
| At 4 years | 3.99 (2.39, 6.34) | 5.52 (3.23, 9.03) | 4.90 (2.87, 8.00) |
| At 10 years | 2.79 (1.76, 4.25) | 3.47 (2.13, 5.41) | 3.21 (1.97, 5.00) |
| *Odds of psychosis in stroke* |  |  |  |
| At 4 years | 3.99 (2.39, 6.34) | 5.14 (2.99, 8.46) | 4.41 (2.57, 7.26) |
| At 10 years | 2.79 (1.76, 4.25) | 3.30 (2.03, 5.17) | 2.94 (1.81, 4.60) |

OR = Odds Ratio. All models were significant at the $p < 0.001$ level. Model 1: adjusted for age, sex, ethnicity, quintile of net financial wealth + participant (frailty). Model 2: adjusted for age, sex, ethnicity, quintile of net financial wealth, smoking status, level of alcohol use in the past 12 months, level of vigorous physical activity + participant (frailty). Adjusted models are reported following multiple imputation of missing baseline covariates.

The cumulative incidence of first-reported stroke, stratified by psychosis status and up to 10 years, is shown in Figure 1. By 10 years, the probability of reporting an incident first stroke in the psychosis group was 21.4% (95% CI, 12.1 to 29.6) compared to 8.3% (95% CI, 7.8 to 8.8) in those without psychosis (absolute difference: 13.1%, 95% CI, 20.8 to 4.3, log rank test, *p* <0.001), equating to an overall unadjusted HR of 2.87 (95% CI, 1.91 to 4.29) (Table 3). When adjusted for baseline demographic covariates, the HR increased to 3.87 (95% CI, 2.45 to 6.11) and when additionally adjusted for baseline health behaviour covariates (smoking status, level of vigorous physical activity and level of alcohol use) the HR reduced slightly to 3.63, with a similar 95% CI (2.52 to 5.87).

**Table 3.** Hazard ratios estimates for stroke risk after psychosis and psychosis risk after stroke.

|  | *Unadjusted*<br>*HR (95% CI)* | *Adjusted, model 1*<br>*HR (95% CI)* | *Adjusted, model 2*<br>*HR (95% CI)* |
| --- | --- | --- | --- |
| Stroke risk after psychosis | 2.87 (1.91, 4.29) | 3.87 (2.45, 6.11) | 3.63 (2.25, 5.87) |
| Psychosis risk after stroke | 3.08 (1.99, 4.77) | 6.06 (3.01, 12.2) | 5.81 (2.89, 11.7) |

<sup>1</sup> HR = Hazard Ratio. All models were significant at the $p < 0.001$ level. Model 1: adjusted for age, sex, ethnicity, quintile of net financial wealth + participant (frailty). Model 2: adjusted for age, sex, ethnicity, quintile of net financial wealth, smoking status, level of alcohol use in the past 12 months, level of vigorous physical activity + participant (frailty). Adjusted models are reported following multiple imputation of missing baseline covariates.

**Figure 1.**
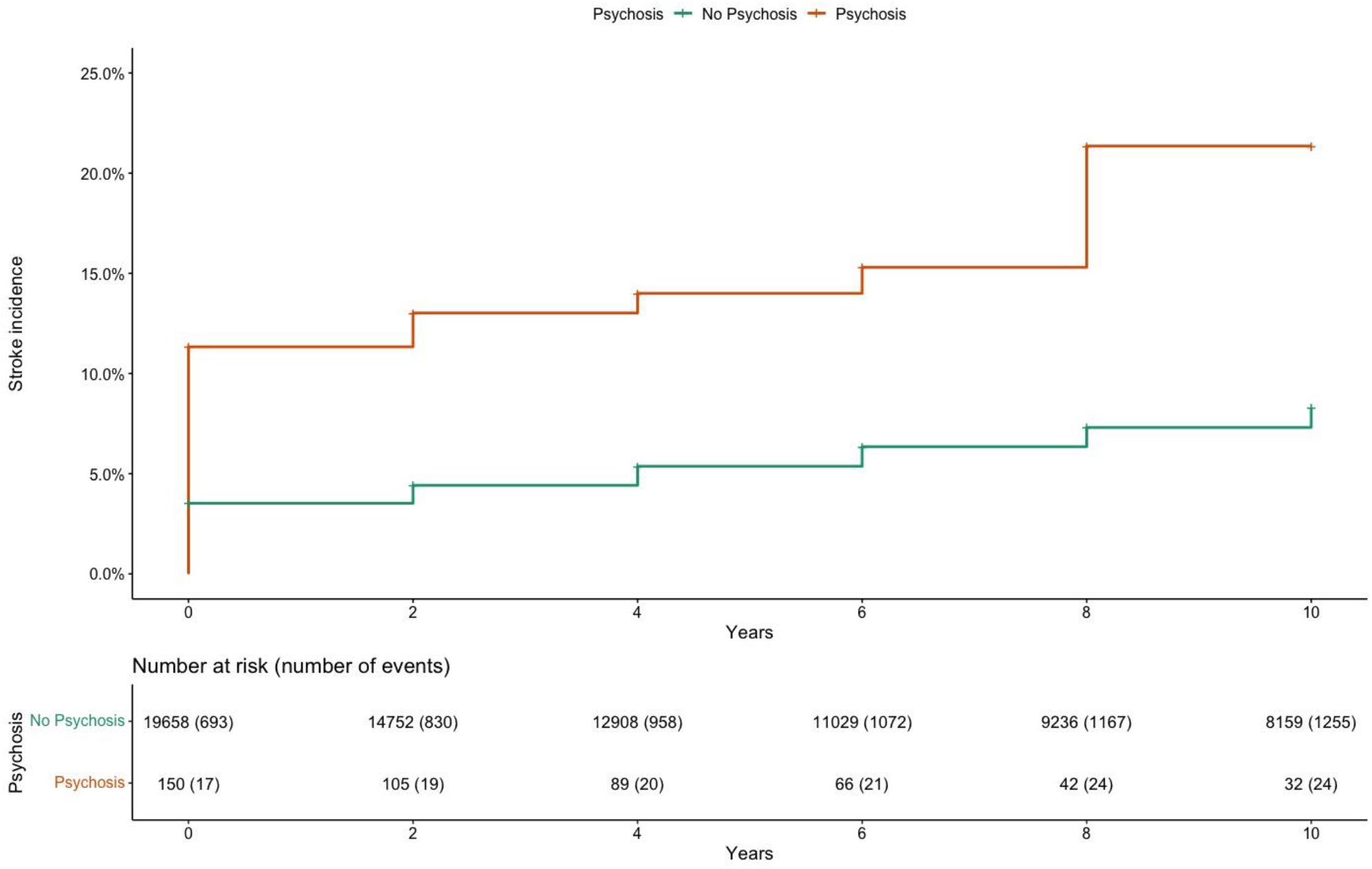
The cumulative incidence of stroke in participants reporting and not reporting psychosis. Log rank test p=<0.001.

### Psychosis risk after stroke

After 4 years of follow-up, and following adjustment for baseline demographic covariates, stroke was associated with increased odds of reporting psychosis compared to people without a stroke (adjusted OR, 5.14, 95% CI, 2.99 to 8.46). When additionally adjusted for baseline health behaviour covariates, the OR reduced to 4.41 (95% CI, 2.57 to 7.26). By 10 years, the ORs had reduced, but continued to indicate that stroke was associated with psychosis (fully adjusted OR, 2.94, 95% CI, 1.81 to 4.60) (Table 2).

The cumulative incidence of first-reported psychosis, stratified by stroke status and up to 10 years, is shown in Figure 2. By 10 years, the probability of reporting incident psychosis in the stroke group was 2.3% (95% CI, 1.4 to 3.2) compared to 0.9% (95% CI, 0.7 to 1.1) in those without stroke (absolute difference: 1.4%, 95% CI, 0.7 to 2.1, log rank test *p*=<0.001), equating to an unadjusted HR of 3.08 (95% CI, 1.99 to 4.77) (Table 3). When adjusted for baseline demographic covariates, the HR increased to 6.06 (95% CI, 3.01 to 12.20), and remained similar when additionally adjusting for baseline health behaviour covariates (5.81, 95% CI, 2.89 to 11.70).

**Figure 2.**
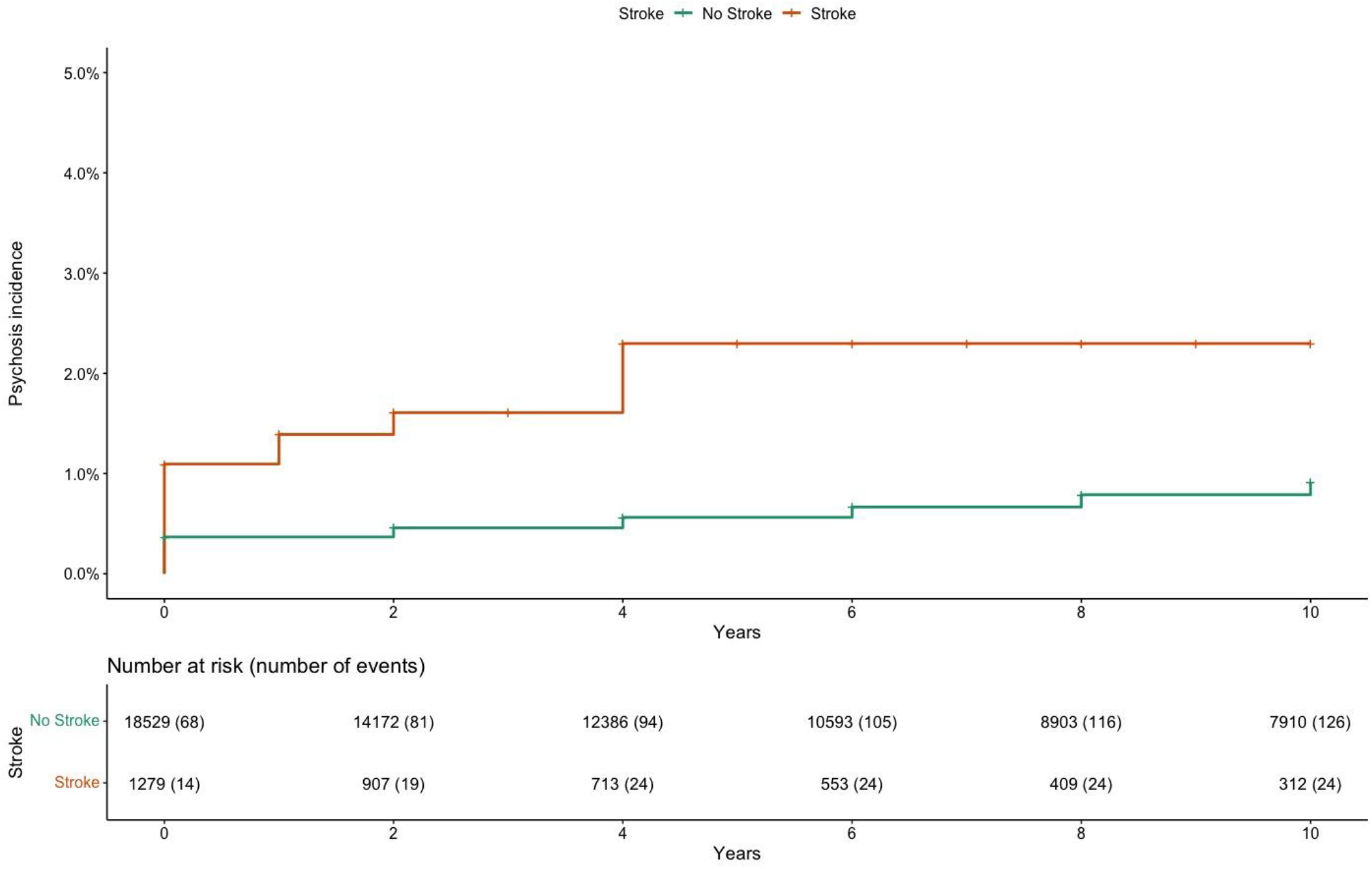
The cumulative incidence of psychosis in those reporting and not reporting stroke. Log rank test p=<0.001.

ORs and HRs from complete-case analysis were broadly similar to those obtained following multiple imputation but tended to result in slightly smaller estimates (Supplementary Tables 3-4).

## Discussion

We report increased odds and hazards of stroke in people with psychosis (and vice versa) over 10 years of follow-up in a representative longitudinal study of adults aged 50 years and over in England. Stroke was an independent predictor of psychosis, and psychosis an independent predictor of stroke, over and above the effects of covariates in adjusted analyses. Stroke-first individuals had a higher prevalence of psychosis at baseline and a higher risk of subsequent psychosis, and psychosis-first individuals showed a similar pattern with regard to later stroke occurrence. However, the risks were not symmetrical: there was a greater risk of psychosis after stroke (fully adjusted HR, 5.81; 95% CI, 2.89 to 11.70) than stroke after psychosis (fully adjusted HR, 3.63; 95% CI, 2.25 to 5.87). These results contribute to our understanding of stroke-psychosis comorbidity, indicating that each raises the risk of the other and, in the context of an ageing population, it is likely the prevalence of this serious comorbidity will increase over time.

This study has several important strengths, including using stratified sampling to be broadly representative of the population of England aged 50 years and over, stroke reporting having been validated by comparison with medical diagnosis in the Health and Retirement study,^15^ and the ability to examine the occurrence of psychosis in stroke-first participants, and stroke in psychosis-first participants, within the same cohort.

However, we note several important study characteristics that are important when interpreting this study. We note the lower reported population prevalence of psychosis reported in this study (0.7%) compared to a meta-analytic estimate of 1.7% in adults in the same 50 years-and-older age range.^20^ In this study, psychosis was measured by a two-stage screening process: individuals were asked if they had been diagnosed with mental health problems, were given a list of conditions if they concurred, and were coded as having psychosis if they indicated “hallucinations”, “schizophrenia” or “psychosis”. Unlike previous studies that measured psychosis with standardised diagnostic interviews^10^ or relied on clinical records of diagnoses (e.g. ^12^), it is possible that this form of self-report is more likely to under-report psychosis due to stigma, lack of insight, lack of diagnosis, or a combination. Indeed, community rates of psychosis in active case ascertainment studies are 2-3 times higher than the rates of diagnosed psychosis,^21,22^ suggesting a significant proportion of undiagnosed psychotic disorders that would be missed even if self-reporting of diagnosis perfectly reflected rates of diagnosis. In addition, due to the episodic nature of psychosis, we were not able to include additional data integrity measures that we did for stroke – namely, removing any cases of stroke where the person reported a stroke diagnosis at one time point, but did not report it at later time points.

We also note a prevalence of psychosis in stroke population of 1.9% and a prevalence of stroke in psychosis population of 16.0%. This compares to recent estimates of a prevalence of psychosis in the stroke of 3.15% (international) and 1.1% (United Kingdom) and a prevalence of stroke in the psychosis population 3.81% (international) and 3.03% (United Kingdom).^10^ There may be several factors contributing to these differences. One may be the relatively small number of stroke and psychosis cases identified, meaning that population estimates may be affected by relatively small differences in case numbers. The under-reporting of psychosis, as discussed above, is likely to be a factor and may particularly contribute to the much higher estimated prevalence of stroke in psychosis than psychosis in stroke. However, it is also worth noting that the previous estimates are drawn from whole population studies and this study is based on the ELSA cohort that includes adults aged 50 and older only and it is currently not clear to what extent prevalence may differ by age cohort. Younger adults with strokes make up 10-15% of the total stroke population^23^ with the prevalence of strokes rapidly increasing in this age cohort.^24^ Given the typically younger age of onset of psychosis, it is possible that younger individuals with stroke-psychosis may have distinct characteristics from older adults with the same and this remains a topic for further investigation.

We also note that follow-up of the ELSA cohort was completed every two years and this may have missed cases of stroke or psychosis in individuals who developed either condition in the time since their last follow-up but before they died. In addition, it is not clear the extent to which stroke-psychosis comorbidity may have led to individuals being lost to follow-up, leading to selection bias due to increased attrition in this group, or potential interactions where, for example, stroke-related aphasia may have affected the diagnosis or reporting of psychosis.^9^ We used the wave that stroke or psychosis were reported as a proxy for when these conditions first occurred; analyses using dates of onset instead would have been more precise, but these were not available.

One potentially curious feature of the analysis was that risk slightly increased after additionally controlling for several potential demographic and health-related confounders. This effect was largely down to frequency of alcohol consumption where those reporting more frequent consumption of alcohol showed lower risk of stroke and psychosis. This finding, that increased frequency of alcohol consumption is associated with better health in the ELSA cohort, has been the focus of dedicated studies^25,26^ that have indicated that these effects may be explained both by poorer baseline health in non-drinkers and generally lower levels of alcohol consumption by volume in adults as they age reflecting a higher prevalence of ‘light, frequent drinkers’.

There are some additional factors we were not able to examine in this study which are likely to be important in understanding stroke-psychosis comorbidity. Data for antipsychotic prescribing were not available at baseline and there is evidence that certain antipsychotic medications may increase the risk of mortality and further stroke morbidity in those with stroke and psychosis.^27^ In addition, data on stroke type, severity and/or location were not available, meaning it was not possible to examine the bidirectional association between stroke characteristics and psychosis. As our focus was on stroke-psychosis comorbidity, we did not study the incidence of other cardiovascular conditions and diseases (e.g., hypertension, myocardial infarction) which also play a role in stroke risk and could represent competing risks. Whether there are shared genetic pathways to stroke and psychosis also remains unexplored.

In conclusion, we report that there is a bidirectional increase in risk for stroke-psychosis comorbidity. There was a greater risk of psychosis after stroke than stroke after psychosis. However, due to the likely under-reporting of psychosis, there is a pressing need for studies that use gold standard stroke and psychosis diagnostic methods within the same large-scale multi-year follow-up study to examine this issue in more detail.

## Data Availability

English Longitudinal Study of Ageing (ELSA) data are publicly accessible; data were accessed via UK Data Service (project ID: 222747). Data cannot be shared by the authors of this article but can be obtained directly from the UK Data Service (https://ukdataservice.ac.uk/).

## Acknowledgements

The English Longitudinal Study of Ageing was developed by a team of researchers based at University College London, NatCen Social Research, the Institute for Fiscal Studies, the University of Manchester and the University of East Anglia. The data were collected by NatCen Social Research. The funding is currently provided by the National Institute on Aging in the US, and a consortium of UK government departments coordinated by the National Institute for Health Research. Funding has also been received by the Economic and Social Research Council.

