## Supplementary Material for "Longitudinal associations between stroke and psychosis: a population-based study"

|  | *No Stroke or Psychosis* | *Psychosis only* | *Stroke and Psychosis* | *Stroke only* |
| --- | --- | --- | --- | --- |
| N | 18,403 | 126 | 24 | 1,255 |
| Stroke |  |  |  |  |
| Age at first stroke, median (IQR) | - | - | 66 (58, 79) | 69 (59, 77) |
| Unknown | *-* | *-* | 2 | 114 |
| Stroke recurrences, mean (SD) | - | - | 2.2 (3.0) | 1.1 (1.5) |
| Stroke recurrences (if >0), mean (SD) | - | - | 3.1 (3.2) | 2.0 (1.5) |
| Unknown | *-* | *-* | 7 | 543 |
| Ever-reported psychiatric diagnoses |  |  |  |  |
| Depression, n (%) | 1,733 (9.4%) | 111 (88.1%) | 20 (83.3%) | 145 (11.6%) |
| Anxiety, n (%) | 1,496 (8.1%) | 104 (82.5%) | 18 (75.0%) | 129 (10.3%) |

### Supplementary Table 1. Stroke status and ever-reported psychiatric diagnoses for sample.

| *Characteristic* | *No Stroke or Psychosis* | *Psychosis only* | *Stroke and Psychosis* | *Stroke only* |
| --- | --- | --- | --- | --- |
| N | 18,403 | 126 | 24 | 1,255 |
| Number of waves participated^1^ | 4.0 (2.0, 7.0) | 5.0 (3.0, 7.0) | 6.0 (3.0, 8.0) | 4.0 (2.0, 6.0) |
| Participated in all waves, n (%) | 3,060 (17%) | 20 (16%) | 2 (8.3%) | 143 (11%) |
| Died before Wave 6, n (%) | 2,293 (12%) | 12 (9.5%) | 5 (21%) | 409 (33%) |

*^1^* Median (IQR)

### Supplementary Table 2. Study participation across stroke and psychosis groups.

|  | *Unadjusted* | *Adjusted, model 1* | *Adjusted, model 2* |
| --- | --- | --- | --- |
|  | OR (95% CI) | OR (95% CI) | OR (95% CI) |
| Odds of stroke in psychosis |  |  |  |
| At 4 years | 3.99 (2.39, 6.34) | 5.00 (2.84, 8.37) | 3.84 (1.99, 6.86) |
| At 10 years | 2.79 (1.76, 4.25) | 3.21 (1.93, 5.10) | 2.62 (1.48, 4.38) |
| Odds of psychosis in stroke |  |  |  |
| At 4 years | 3.99 (2.39, 6.34) | 4.73 (2.67, 7.97) | 3.45 (1.77, 6.24) |
| At 10 years | 2.79 (1.76, 4.25) | 3.12 (1.88, 4.98) | 2.45 (1.38, 4.11) |

*^1^* OR = Odds Ratio. All models were significant at the p<0.001 level. Model 1: adjusted for age, sex, ethnicity, quintile of net financial wealth + participant (frailty). Model 2: adjusted for age, sex, ethnicity, quintile of net financial wealth, smoking status, level of alcohol use in the past 12 months, level of vigorous physical activity + participant (frailty).

### Supplementary Table 3. Odds ratios estimates for stroke risk after psychosis and psychosis risk after stroke (complete-case).

|  | *Unadjusted* | *Adjusted, model 1* | *Adjusted, model 2* |
| --- | --- | --- | --- |
|  | HR (95% CI) | HR (95% CI) | HR (95% CI) |
| Stroke risk after psychosis | 2.87 (1.91, 4.29) | 3.44 (2.21, 5.36) | 3.18 (1.80, 5.61) |
| Psychosis risk after stroke | 3.08 (1.99, 4.77) | 4.65 (2.44, 8.86) | 5.01 (2.19, 11.5) |

*^1^* HR = Hazard Ratio. All models were significant at the p<0.001 level. Model 1: adjusted for age, sex, ethnicity, quintile of net financial wealth + participant (frailty). Model 2: adjusted for age, sex, ethnicity, quintile of net financial wealth, smoking status, level of alcohol use in the past 12 months, level of vigorous physical activity + participant (frailty).

### Supplementary Table 4. Hazard ratios estimates for stroke risk after psychosis and psychosis risk after stroke (complete-case).
